# Possibilities of exponential or Sigmoid growth of Covid19 data in different states of India

**DOI:** 10.1101/2020.04.10.20060442

**Authors:** Supriya Mondal, Sabyasachi Ghosh

## Abstract

We have attempted to understand existing covid19 data of India, where growth of total and new cases with time in different states are kept as focal points. Identifying the last trend of exponential growth, mainly noticed in month of March, we have zoomed in its disaster possibilities by straight forward extrapolation of exponential growth. As a hopeful extrapolation, the existing data might be considered low time-axis values of Sigmoid-type function, whose growth might be saturated to values of 10^4^ or 10^5^. To fulfill this expectation, a turning from increasing to decreasing trend in new case data should be noticed around April-May, which definitely demand extension of present lock-down with additional interventions.

## 1 Introduction

The Covid19 spreading is started from wuhan, China on December 2019 and then within a three months time period this infection spreads almost throughout the world. Therefore, this covid19 infection is declared as pandemic disease by World Health Organization (WHO) [1]. In India, first entry from Wuhan,China is noticed on 30th of January, 2020 [2]. It remain quite gentle up to February and limited within Kerala but from beginning of March, it spread to other states and union territories of India. During the month of March the number of covid19 cases get an exponential growth tendency. On the identification of exponential growth trend and its possible deviation via Sigmoid function [3] for India data has been attempted in our earlier work [4]. Present work can be considered as continuation of earlier investigation. We focus here on the covid19 data of individual states and union territories of India.

In recent time, epidemic dynamics of covid19 have been described in Refs [5, 6] through the Sigmoid-type logistic function. Similar kind of phenomenological investigation is initiated by Ref. [7] through renormalization group inspired logistic function. The epidemiology study on covid19 data of India is started by few Refs. [8, 4], which should be rapidly grown by other theoretical groups for better and critical understanding. In that context, present work have attempted to provide initial quantitative documentation, which probably invite more future works.

The article is organized as follows. Next section (2), we have addressed the mathematical framework part of simple form of exponential and Sigmoid-type function. Then results of different states are analyzed through graphical representations in Sec. (3). Brief bullet points of different states are mentioned in their respective subsections. At the end, we have summarized the study with extracting interpretation in Sec. (4).

## 2 Mathematical Framework

We have identified the exponential growth in time profile of total cases for different states, where some of them follow it in entire time zone (approximately) and remaining of them follow it in particular time zone(s). For later case, we will take interest on last part of exponential trend and its corresponding growth parameter *λ*. Simple form of exponential growth is based on the fact that rate of increment is proportional to total cases, i.e.

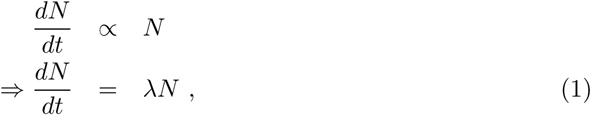

where proportional constant *λ* is called growth parameter. The integral form of Eq. (1) is

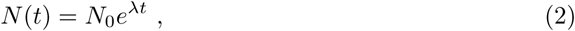

where *N*_0_ is initial values of +ve cases. Eq. (1) can alternatively realized as no. of new cases (*dN*) = *λ*× no. of total cases (*N*). So if we find that total number of cases follow Eq. (2) in entire *t*-axis or some part of *t*-axis, then proportional relation between new cases and total cases can be seen in that *t* range(s). Fig. (1) shows the exponential fitting of either entire *t*-axis or some part of *t*-axis, where we will take interest on last part of exponential slope. Since *y*-axes in Fig. (1) are plotted in logarithmic scale, so their entire/last slope will give us values of *λ*, marked in their respective legend boxes.

**Figure 1:**
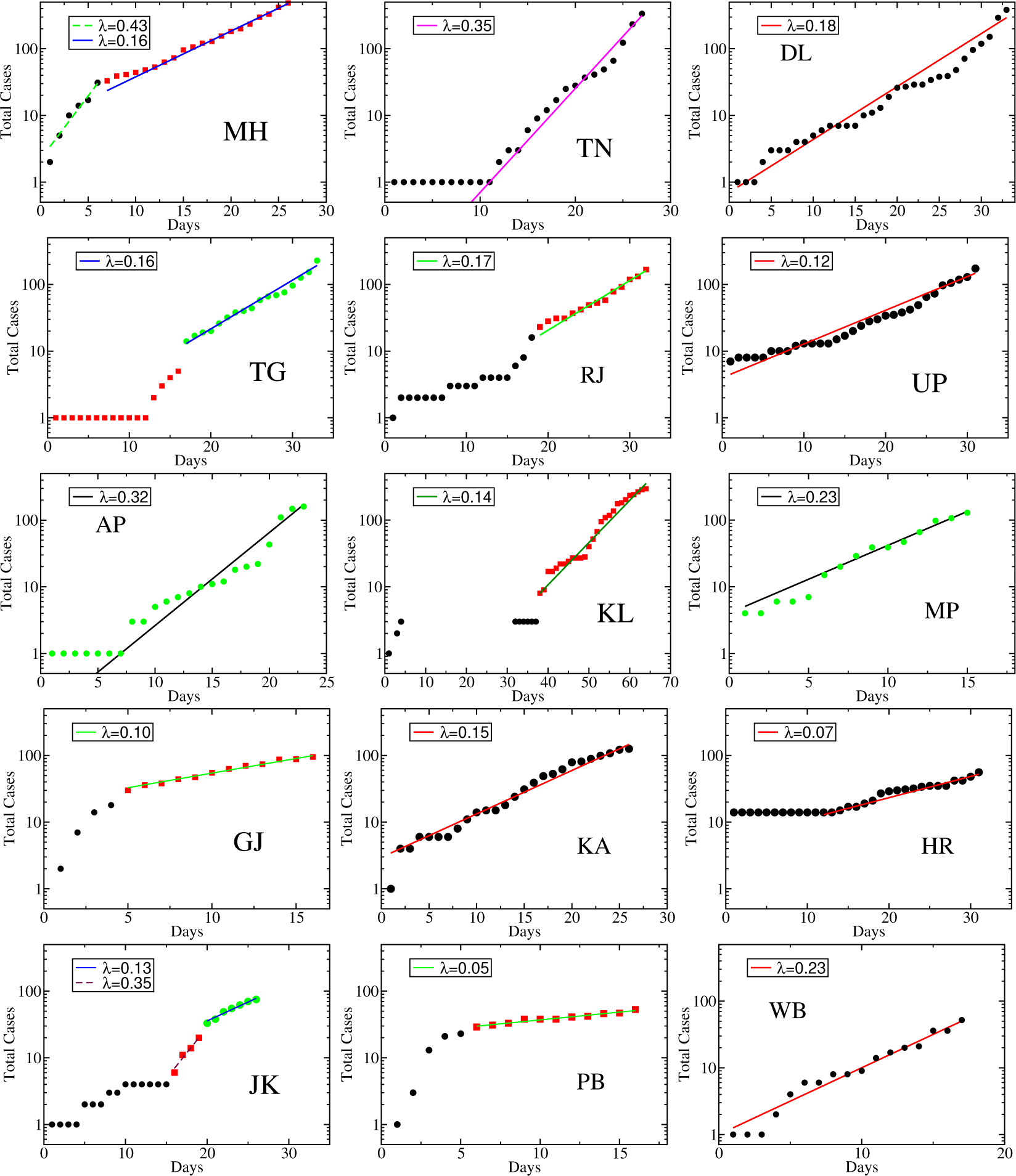
The exponential fitting of total cases vs time (in days) for different states and their growth parameter *λ*’s are written in legend box.

To prevent this massive infection spread, the deviation of exponential function is the option. In logarithmic scale, we can imagine a better situation just by decreasing the slopes or *λ*, and at the end, making total cases be started to constant values. One of the example of this kind of curve is Sigmoid curve [3], which can be realized in terms of a transformed exponential growth. Sigmoid function will also have an initial exponential growth, which will be flattened due to an extra time dependent fraction

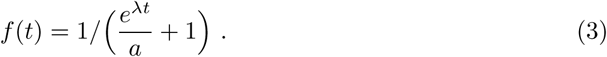

Here, *a* is very important parameter, which fix the maximum number of cases *N*_*max*_ = *N*_0_×*a*, where the sigmoid curve will saturate. So in terms of Eqs. (2) and (3), we can write Sigmoid function as

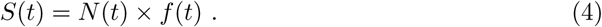

Similar to rate Eq. (1), time derivative of Sigmoid function is

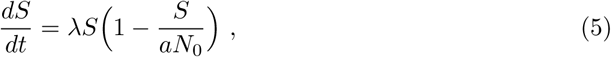

which shows that number of new cases (*dS*) has two parts - one is proportionally growing with total cases (i.e. ∝ *S*) and another is decaying with ∝ *S*^2^*/N*_*max*_. In small values of *t*, Eqs. (5), (4) will convert to Eqs. (1), (2). It means that Sigmoid function will behave as exponential function in low *t* range but at high *t*, it will saturate to a maximum values (*N*_*max*_). On the other hand, its derivative or slope (no of new cases) first increase, then decrease after showing a peak. By taking a guess value of *N*_*max*_ = 10^4^ and 10^5^, we have drawn different possible Sigmoid functions and their derivative (new cases) for different states, which are addressed in next section.

## 3 Results of Different States

We have first taken the raw data from Ref. [2], then we have make data file for different states having columns - days, new cases and total cases. We have started day one from when non-zero number of covid19 +ve case is detected. Among 28 states and 8 union territories, we have considered only 15, whose total number of covid19 +ve cases are quite larger (> 40 on 3rd April). They are (1) Maharashtra (MH), (2) Tamil Nadu (TN), (3) Delhi (DL), (4) Telangana (TG), (5) Kerala (KL), (6) Uttar Pradesh (UP), (7) Andhra Pradesh (AP), (8) Rajasthan (RJ), (9) Madhya Pradesh (MP), (10) Karnatak (KA), (11) Gujrat (GJ), (12) Haryana (HR), (13) Jammu & Kashmir (JK), (14) Punjab (PB), and (15) West Bengal (WB). We have gone through them one by one with graphical represenations and discussions.

### 3.1 Maharashtra (MH)

According to present data, highest number of +ve cases has been noticed in Maharashtra (MH). We noticed in MH-panel of Fig. (1), that last slope parameter is *λ* = 0.16, which was previously *λ* = 0.43. This larger to lower exponential transformation is definitely a good signal for MH but further extension of last exponential growth is so dangerous that it can cover full population of MH within the end of June, 2020. In the upper left panel of Fig. (2), the pink dotted line represent that exponential growth, where number of population (NoP) in MH is marked by red arrow. However, proper interventions like lock-down, maintaining self precautions can transform the exponential to Sigmoid-type profile. Considering two guess values *N*_*max*_ = 10^4^, 10^5^, where total cases might be saturated due to undertaking interventions, we can get two Sigmoid functions *S*_1_ and *S*_2_ respectively, shown by blue dash and black solid lines. Time derivative of exponential function and Sigmoid functions will give corresponding new case predictions, which is shown in lower left panel of Fig. (2) with same line styles as considered in upper panel for total case plots. New case data of MH is also included. They are quite fluctuating but matched roughly with low *t* range of exponential and Sigmoid functions. For predicted hopeful functions *S*_1_ and *S*_2_, the turning of new cases from increasing to decreasing trend has to be seen around end of April and mid of May. Number of Beds (NoB) in Govt. hospitals in different states are documented in the Ref. [9]. For MH, this number is shown by green dash line in left upper and lower panels of Fig. (2).

In brief, our bullet point outcomes of MH are as follows.

- Starting date is 9th March (2020)
- Last slope parameter of log-scale total cases is *λ* = 0.16.
- Continuation of exponential growth can cover NoP within the end of June (2020).
- *S*_2_ > NoB > *S*_1_.
- Peak of new cases for *S*_1_,_2_ are expected around end of April, mid of May (2020).

**Figure 2:**
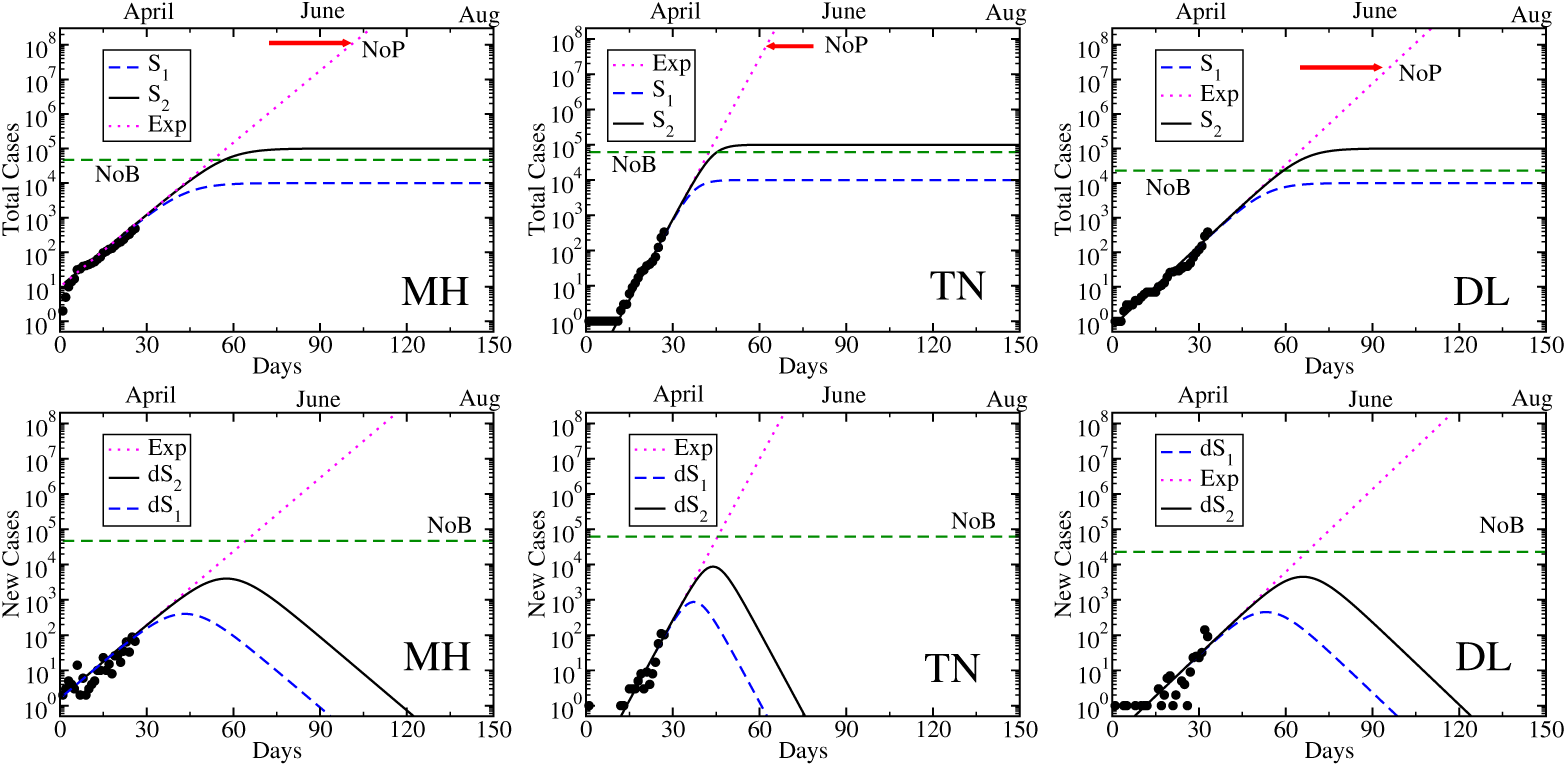
Total cases (upper), new cases (lower) of MH (left), TN (middle) and DL (right), where actual data (black circles), exponential (pink dotted line) and *S*_1_ (blue dash line), *S*_2_ (black solid line) are plotted with marking NoP (red arrow) and NoB (green dash line).

**Figure 3:**
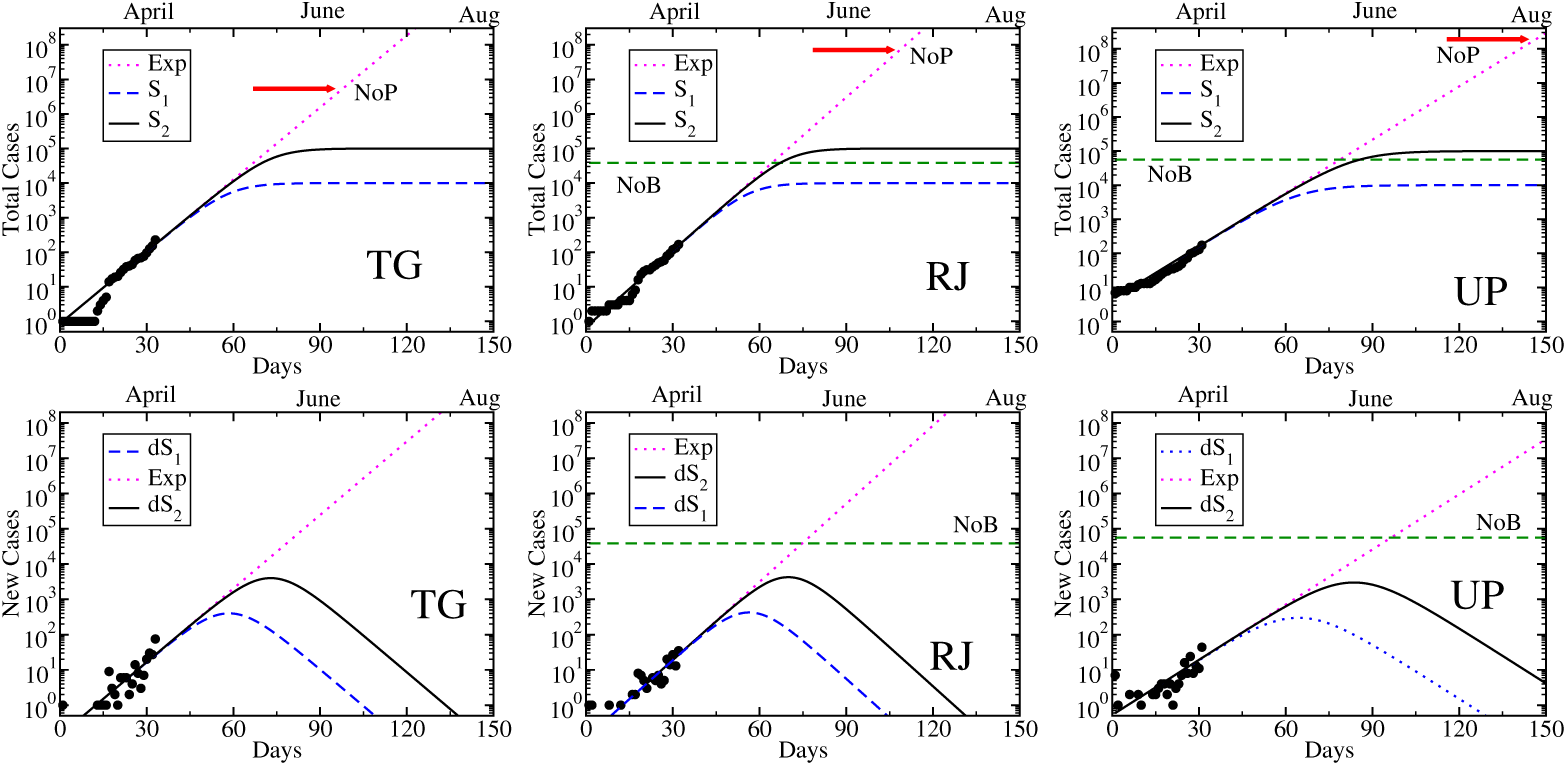
Same as Fig. (2) for TG (left), RJ (middle) and UP (right).

**Figure 4:**
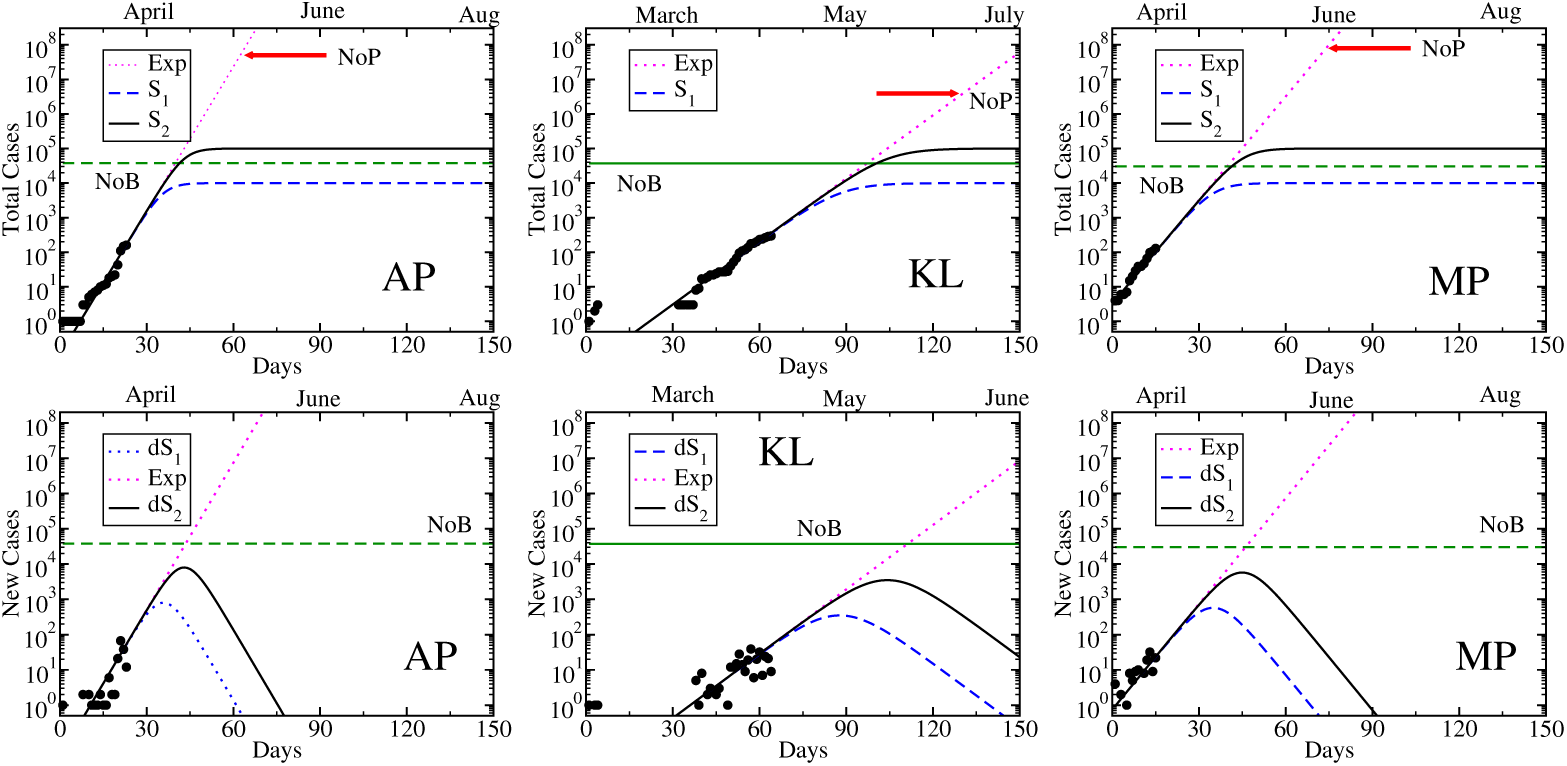
Same as Fig. (2) for AP (left), KL (middle) and MP (right).

**Figure 5:**
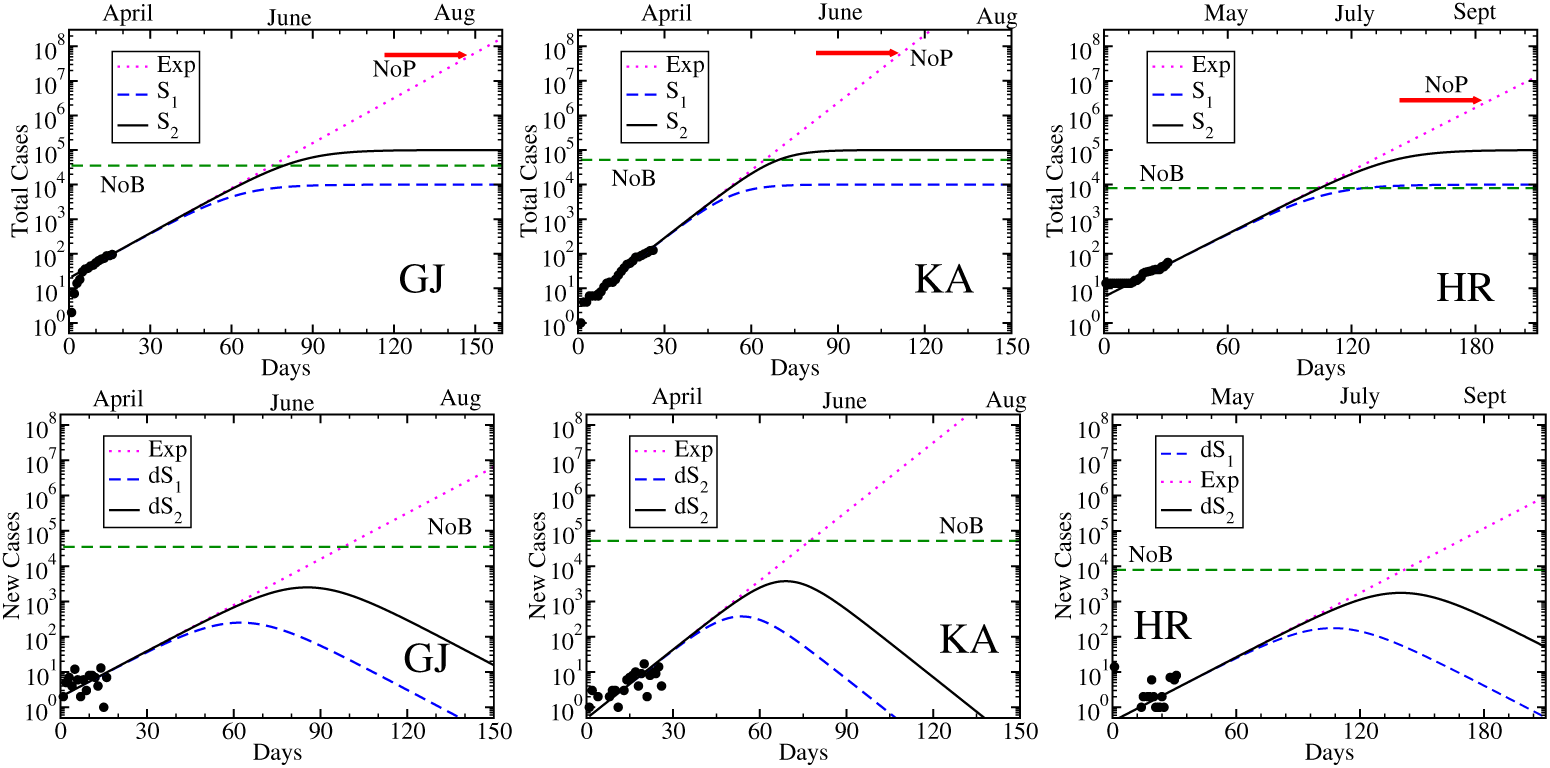
Same as Fig. (2) for GJ (left), KA (middle) and HR (right).

**Figure 6:**
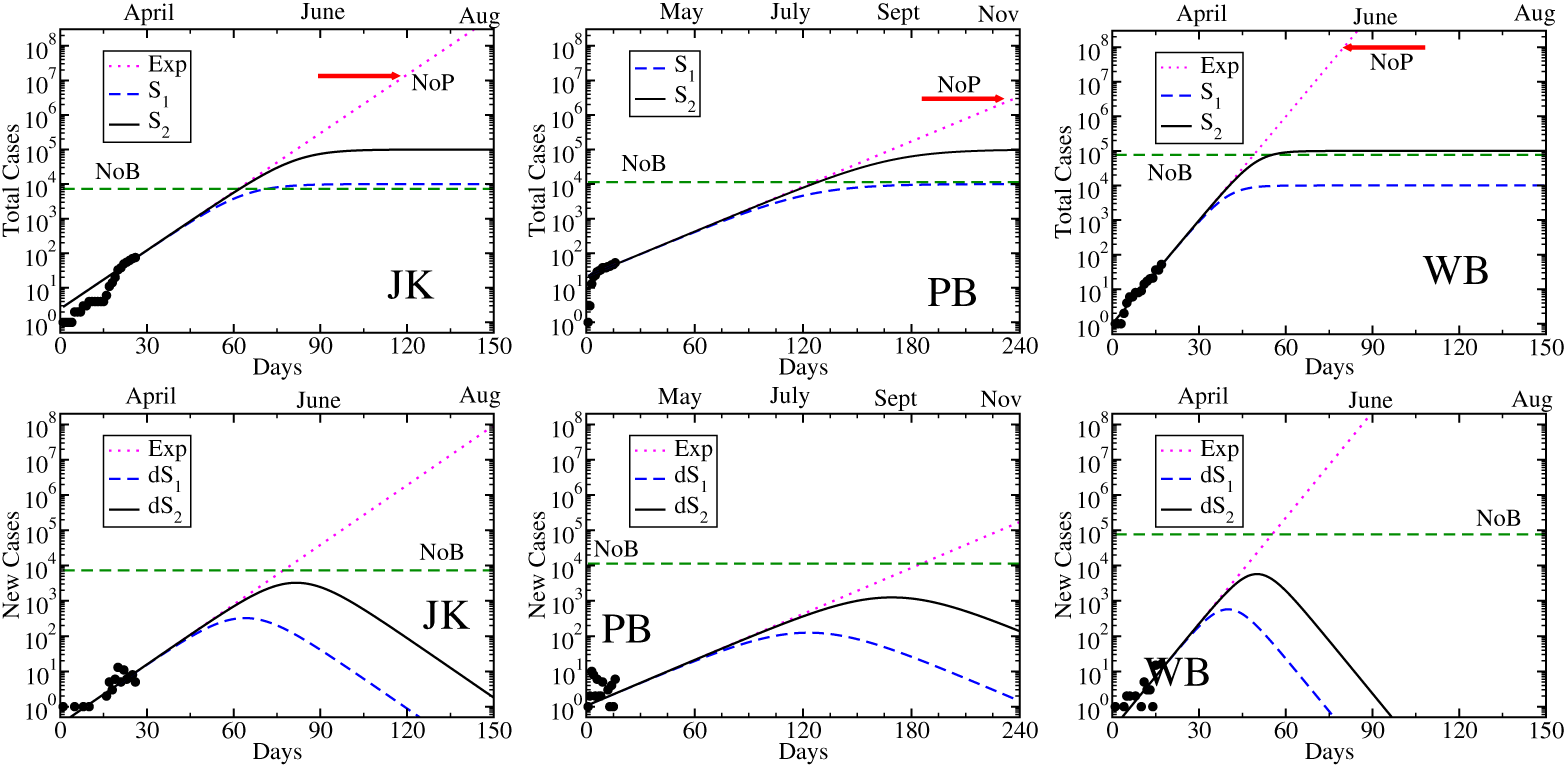
Same as Fig. (2) for JK (left), PB (middle) and WB (right).

For remaining states and union territories, we have followed similar kind of sketching, hence instead of elaborate discussions of their corresponding graphs, main bullet points are highlighted in their respective subsections, addressed below.

### 3.2 Tamil Nadu (TN)

- Starting date is 7th March (2020)
- Last slope parameter of log-scale total cases is *λ* = 0.35.
- Continuation of exponential growth can cover NoP within the mid of May.
- *S*_2_ > NoB > *S*_1_.
- Peak of new cases for *S*_1_,_2_ are expected around mid of April, end of April.

### 3.3 Delhi (DL)

- Starting date is 2nd March (2020)
- Last slope parameter of log-scale total cases is *λ* = 0.18.
- Continuation of exponential growth can cover NoP within the mid of June.
- *S*_2_ > NoB > *S*_1_.
- Peak of new cases for *S*_1_,_2_ are expected around end of April, mid of April.

### 3.4 Telangana (TG)

- Starting date is 2nd March (2020)
- Last slope parameter of log-scale total cases is *λ* = 0.16.
- Continuation of exponential growth can cover NoP within the mid of June.
- Peak of new cases for *S*_1_,_2_ are expected around beginning of May, mid of May.

### 3.5 Rajasthan (RJ)

- Starting date is 3rd March (2020)
- Last slope parameter of log-scale total cases is *λ* = 0.17.
- Continuation of exponential growth can cover NoP within the end of June.
- *S*_2_ > NoB > *S*_1_.
- Peak of new cases for *S*_1_,_2_ are expected around end of April, mid of May.

### 3.6 Uttar Pradesh (UP)

- Starting date is 4th March (2020)
- Last slope parameter of log-scale total cases is *λ* = 0.12.
- Continuation of exponential growth can cover NoP within the end of August.
- *S*_2_ > NoB > *S*_1_.
- Peak of new cases for *S*_1_,_2_ are expected around beginning of May, end of May.

### 3.7 Andhra Pradesh (AP)

- Starting date is 12th March (2020)
- Last slope parameter of log-scale total cases is *λ* = 0.32.
- Continuation of exponential growth can cover NoP within the mid of May.
- *S*_2_ > NoB > *S*_1_.
- Peak of new cases for *S*_1_,_2_ are expected around mid of April, end of April.

### 3.8 Kerala (KL)

- Starting date is 30th January (2020)
- Last slope parameter of log-scale total cases is *λ* = 0.14.
- Continuation of exponential growth can cover NoP within the mid of June.
- *S*_2_ > NoB > *S*_1_.
- Peak of new cases for *S*_1_,_2_ are expected around end of April, mid of May.

### 3.9 Madhya Pradesh (MP)

- Starting date is 20th March (2020)
- Last slope parameter of log-scale total cases is *λ* = 0.23.
- Continuation of exponential growth can cover NoP within the End of June.
- *S*_2_ > NoB > *S*_1_.
- Peak of new cases for *S*_1_,_2_ are expected around End of April, beginning of May.

### 3.10 Gujarat (GJ)

- Starting date is 19th March (2020)
- Last slope parameter of log-scale total cases is *λ* = 0.10.
- Continuation of exponential growth can cover NoP within the mid of August.
- *S*_2_ > NoB > *S*_1_.
- Peak of new cases for *S*_1_,_2_ are expected around mid of May, beginning of June.

### 3.11 Karnatak (KA)

- Starting date is 9th March (2020)
- Last slope parameter of log-scale total cases is *λ* = 0.15.
- Continuation of exponential growth can cover NoP within the beginning of July.
- *S*_2_ > NoB > *S*_1_.

Peak of new cases for *S*_1_,_2_ are expected around beginning of May, mid of May.

### 3.12 Haryana (HR)

- Starting date is 4th March (2020)
- Last slope parameter of log-scale total cases is *λ* = 0.07.
- Continuation of exponential growth can cover NoP within the beginning of September.
- *S*_2_ > *S*_1_ > NoB.
- Peak of new cases for *S*_1_,_2_ are expected around Mid of June, end of August.

### 3.13 Jammu Kashmir (JK)

- Starting date is 9th March (2020)
- Last slope parameter of log-scale total cases is *λ* = 0.13.
- Continuation of exponential growth can cover NoP within the beginning of July.
- *S*_2_ > *S*_1_ > NoB.
- Peak of new cases for *S*_1_,_2_ are expected around mid of May, beginning of June.

### 3.14 Punjab (PB)

- Starting date is 19th March (2020)
- Last slope parameter of log-scale total cases is *λ* = 0.05.
- Continuation of exponential growth can cover NoP within the beginning of November.
- *S*_2_ > NoB > *S*_1_.
- Peak of new cases for *S*_1_,_2_ are expected around beginning of July, end of August.

### 3.15 West Bengal (WB)

- Starting date is 17th March (2020)
- Last slope parameter of log-scale total cases is *λ* = 0.23.
- Continuation of exponential growth can cover NoP within the beginning of June.
- *S*_2_ > NoB > *S*_1_.
- Peak of new cases for *S*_1_,_2_ are expected around end of April, beginning of May.

## 4 Summary and Interpretation

Present investigation has attempted to provide the sketch of covid19 spreading with days in different states and union territories of India. Among the 28 states and 8 union territories, we have considered only 15, whose total number of covid19 +ve cases are quite larger. Except KL remaining of them have been detected first covid19 positive cases from the beginning of March. Using this one month existing data of total and new cases for different states, we have attempted to visualize their future trends. Identifying the last trend of exponential growth of different states, we have listed their slope parameters (*λ*) from their respective time dependent total case plot. Though we have arranged our descriptions from MH to WB, based on total no of cases in an ascending order but they don’t have same ascending order in their slope parameters, which is very important quantity for future spreading. In that context, TN, AP, MP, WB, having higher slope parameters, might face rapid growth of positive cases, while states having low slope parameter, like GJ, HR, PB might face slow growth. To zoom in the disaster side of this exponential growth we have extrapolated the total cases based on the last slope parameters and we estimated approximate months when entire population of the states might be infected. Due to the high slope parameter, TN and AP might reach this picture within May while WB, DL, TG, KL, MH, RJ, MP might reach on June, then UP, GJ on August; HR on September; PB on November. This order probably reflect the ranking of states, based on interventions, maintained by state residents.

After the extrapolation of disaster side of last growth trend, a hopeful possibility is sketched via Sigmoid function, which can also be extrapolated from same existing data. We consider the two guess values 10^4^ and 10^5^, where total cases might be saturated due to undertaking interventions. Then we have plotted two Sigmoid-type profile in time axis. With respect to existing data for available beds in Govt. hospitals, JK, HR, PB are in little high risk zone in comparison to others as the rough values of their no. of beds remain lower than saturation number of Sigmoid functions. Interestingly, their high risk might be reduced by their ongoing undertaking interventions, for which their last trend of exponential growth have very small slope parameters. On the other hand WB, TN, AP due to their large values of Govt. hospital beds, seems to be in low risk zone but in the same time we should not ignore their high risk possibility because of their large slope parameter, noticed in their data. As we know that Sigmoid-type profile in total cases can create a peak structure in new case profile, which means that after a certain time, increasing trend of new cases can be converted to decreasing trend. To get saturation in total cases of 10^4^ and 10^5^, maximum states have to face the turning from increasing to decreasing trend in new cases around April or May. However, this time for HR and PB might be relaxed to June or August due to their lower slope parameters.

During the ongoing lock-down from 25th March, imposed by India Govt., present data does not face any noticeable deviation from exponential growth, whose origin is probably linked with some accidental events before 22nd March or latter. This fact is really matter of critical research but a straight forward extension of lock-down definitely a hope for getting deviation from exponential growth. In that case, a turning possibility in new cases at the end of the April or May might be expected. Along with the extension of present lockdown, compensating loop-holes in interventions is probably very important task and more additional interventions might be necessary if required.

We believe that present investigation might play as an important initial portrait to understand current epidemiology status in different states and union territories of India. It might be helpful for further critical investigations, through which we can get better understanding than now. It desperately invites to different theoretical minds to probe this epidemiology picture in more deep understanding, whose delayed response might be too late for society.

## Data Availability

cited in references

## Acknowledgment

SM and SG thank to their daughter Adrika Ghosh for allowing time for this investigation during lock-down period.

